# Clinical Performance of the cobas Liat SARS-CoV-2 & Influenza AB for the Detection of SARS-CoV-2 in Nasal Samples

**DOI:** 10.1101/2022.01.07.22268874

**Authors:** Yusaku Akashi, Michiko Horie, Junichi Kiyotaki, Yuto Takeuchi, Kenichi Togashi, Yuki Adachi, Atsuo Ueda, Shigeyuki Notake, Koji Nakamura, Norihiko Terada, Yoko Kurihara, Yoshihiko Kiyasu, Hiromichi Suzuki

**Author notes:** **Authors’ E-mail addresses** Michiko Horie,; Junichi Kiyotaki,; Yuto Takeuchi,; Kenichi Togashi,; Yuki Adachi,; Atsuo Ueda,; Shigeyuki Notake,; Koji Nakamura,; Norihiko Terada,; Yoko Kurihara,; Yoshihiko Kiyasu,; Hiromichi Suzuki. **Corresponding author** Yusaku Akashi, M.D., Ph.D., Department of Infectious Diseases, Faculty of Medicine, University of Tsukuba, 1-1-1, Tennodai, Tsukuba, Ibaraki 305-8575, Japan.

## Abstract

**Background and Objective:** Point-of-care type molecular diagnostic tests have been used for detecting SARS-CoV-2, although their clinical utility with nasal samples has yet to be established. This study evaluated the clinical performance of the cobas Liat SARS-CoV-2 & Influenza AB (Liat) in nasal samples.

**Methods:** Nasal and nasopharyngeal samples were collected and were tested using the Liat, the cobas 6800 system and the cobas SARS-CoV-2 & Influenza AB (cobas), and a method developed by National Institute of Infectious Diseases, Japan (NIID).

**Results:** A total of 814 nasal samples were collected. The Liat assay was positive for SARS-CoV-2 in 113 (13.9%). The total, positive, and negative concordance rate between the Liat and cobas/NIID assays were 99.3%/98.4%, 99.1%/100%, and 99.3%/98.2%, respectively. Five samples were positive only using the Liat assay. Their Ct values ranged from 31.9 to 37.2. The Ct values of the Liat assay were significantly lower (p < 0.001) but were correlated (p < 0.001) with those of other molecular assays. In the participants who tested positive for SARS-CoV-2 on the Liat assay using nasopharyngeal samples, 88.2% of their nasal samples also tested positive using the Liat assay.

**Conclusion:** The Liat assay showed high concordance with other molecular assays in nasal samples. Some discordance occurred in samples with Ct values > 30 on the Liat assay.

**Key Points:**

i. The cobas Liat SARS-CoV-2 & Influenza AB assay showed high concordance with other molecular assays in nasal and nasopharyngeal samples, providing results within 20 minutes.
ii. Some discordance occurred in samples with Ct values > 30 on the Liat assay.
iii. The Liat assay may be suitable for use in a variety of clinical situations, primarily where accurate detection of SARS-CoV-2 is necessary.

## 1. Introduction

The pandemic of severe acute coronavirus 2 (SARS-CoV-2) has had a detrimental effect on society globally [1]. The introduction of effective vaccines [2] and treatment [3] were expected to effectively control the pandemic; however, the numbers of new infections and deaths have increased in some countries [1]. Currently, population-based screening and early detection and isolation of infected individuals are still key to effective infection control [4].

The standard method for detecting SARS-CoV-2 is molecular testing due to its high diagnostic performance [5]. However, molecular diagnostics are less widely available and less convenient than antigen testing, and the turnaround time is longer [6]. Several molecular point-of-care tests (POCTs) have been developed [7] and applied in clinical settings to overcome the disadvantages of traditional molecular tests.

The cobas Liat system (Roche Molecular Systems, Inc., Pleasanton, CA, USA) is a small analyzer that automatically performs real-time polymerase chain reaction (PCR) testing using simple procedures and provides results within 20 minutes [8]. The system has been used for point-of-care testing, and Liat assays are available for influenza virus [9], respiratory syncytial virus [10], *Clostridioides difficile* [11], group A *Streptococcus* [12], and SARS-CoV-2 [8]. The Liat system and cobas Liat SARS-CoV-2 & Influenza AB (Liat) has been shown to have high sensitivity for detecting SARS-CoV-2 in nasopharyngeal samples [8], but the diagnostic performance with nasal samples has not been reported. Nasal sample collection is easier and less invasive than the nasopharyngeal sample collection [13]; therefore, the clinical utility of Liat would be greater if it could accurately detect SARS-CoV-2 in nasal samples.

In this study, we prospectively evaluated the diagnostic performance of the Liat SARS-CoV-2 assay using paired samples collected from the anterior nostrils and the nasopharynx of participants. The Liat results of the anterior nasal samples were compared with the results of two other real-time PCR tests.

## 2. Methods and Materials

This study was conducted in a PCR center located in Tsukuba Medical Center Hospital (TMCH) in Japan between July 7 and 29, 2021. The participants were outpatients who were suspected to have SARS-CoV-2 infection due to their symptoms or the history of close contact who underwent in-house PCR testing at the center [14]. They included individuals referred from 51 healthcare facilities and a local healthcare center, and healthcare workers working at TMCH. Clinical information of the participants was routinely recorded along with the in-house PCR. Verbal informed consent was obtained from all participants, and the ethical board of the University of Tsukuba approved the study (approval number: R03-41).

### 2.1 Sample collection

One nasopharyngeal and two nasal samples were collected from each participant. The nasopharyngeal samples were collected as described previously [15]. Nasal samples were collected from the anterior nostrils according to recommended procedures [16]. Briefly, the swab was inserted to a depth of approximately 2 cm and rotated four times against the nasal mucosa. The procedure was repeated in the other nostril using the same swab. One nasal swab sample was used for molecular testing using Liat, and the other nasal swab sample was used for the evaluation of antigen testing in another study. All nasal sample collections for molecular testing followed those for antigen testing.

Both the nasal and nasopharyngeal swab samples were diluted in Universal Transport Medium (UTM™; Copan Diagnostics Inc., Murrieta, CA, USA) and stored at −80°C. The nasopharyngeal samples were frozen after being testing using an in-house PCR test.

### 2.2 PCR testing procedures

After being thawed, all nasal samples underwent PCR testing according to three methods: i) cobas Liat system and cobas Liat SARS-CoV-2 & Influenza AB (Liat), ii) cobas 6800 system and cobas SARS-CoV-2 & Influenza AB (cobas; Roche Molecular Systems, Branchburg, NJ, USA) [17], and iii) a national standard method developed by National Institute of Infectious Diseases (NIID), Japan [18]. If a sample showed positivity on one of the methods and negativity on the other two, they were tested using the Gene Xpert system and the Xpert Xpress SARS-CoV-2 (Xpert Xpress; Cepheid, Sunnyvale, CA, United States) [19].

The nasopharyngeal samples were tested using Liat and the NIID test for the evaluation and were not tested using the cobas system due to an insufficient amount of residual UTM sample. If the Liat and NIID results were discordant, the samples were tested using Xpert Xpress, in a similar manner to the nasal samples.

Liat, cobas, and Xpert Xpress perform sample preparation (purification and the extraction of RNA), real-time PCR, and the detect of the viruses using a fully automated process. For the Liat assay, a total of 200μL of UTM™ sample was loaded into the test cartridge, which was then inserted into the system. The Liat test targets the ORF1a/b and N gene and shows positive results if one or both genes are detected. We re-tested samples if invalid results occurred. For the cobas assay, we used 1 mL of UTM™ sample (400 μL for the analysis and 600 μL for the dead space). The targeted regions were ORF1a/b and E gene, and the cobas test provided separate results for each target. The results of cobas were considered positive for SARS-CoV-2 when samples tested positive for one of the two targets. The Xpert Xpress used 300 μL of UTM™ sample and targeted the E and N2 genes. Similar to cobas, Xpert Xpress showed separate results for each target. All three assays were conducted according to the manufacturer’s instructions.

For the NIID test, purification and RNA extraction were performed using MagNA Pure 96 total NA Isolation Kit and the MagNA Pure 96 Instrument (Roche Molecular Systems, Branchburg, NJ, USA) from 140-μL aliquots of UTM™ sample. The NIID test targets the N2 region, and the equipment used for the RT-PCR included the PCR LightCycler^®^480 Instrument C (Roche Diagnostics International Ltd, Rotkreuz, Switzerland), the QuantiTect^®^ Probe RT-PCR Kit (QIAGEN, Hilden, Germany), and a SARS-CoV-2 positive control (Nihon Gene Research Laboratories, Sendai, Japan). The RT-PCR was performed in duplicate.

The Ct values were automatically calculated after the detection of SARS-CoV-2 for all test methods used in this study.

Apart from detecting SARS-CoV-2, the Liat assay can also detect influenza virus; however, the results have not been re-evaluated and validated by confirmatory molecular assays in this study.

### 2.3 Statistical analyses

The results of the Liat assay were compared with those of the other molecular assays, and the concordance was calculated with 95% confident intervals (CIs), using the Clopper and Pearson method. The Wilcoxon signed-rank test was used to compare the median Ct values. The correlations of the Ct values between two molecular assays were assessed using Pearson’s product-moment correlation coefficient. The statistical analyses were performed using R version 3.14.1 (R Foundation for Statistical Computing, Vienna, Austria), and the figures were created using Python version 3.8.12 (Python Software Foundation, Wilmington, DE, USA).

## 3. Results

The study included a total of 843 participants, of whom 29 provided only nasopharyngeal samples. Therefore, we obtained 814 nasal and 843 nasopharyngeal samples. The Liat assay could not be performed on one nasopharyngeal sample due to the insufficient amount of residual sample (case ID: T0725).

Four nasal samples and twenty-three nasopharyngeal samples exhibited invalid results on the first test of Liat assay. In the second (repeat) test, all four nasal samples tested negative, one nasopharyngeal sample tested positive, and 13 nasopharyngeal samples tested negative. Due to insufficient residual sample volume, the second test could not be performed on one of the nasopharyngeal samples. The nasopharyngeal sample was excluded from the comparative analysis of the Liat and the NIID assay (case ID: T0613). The remaining eight nasopharyngeal samples tested negative following the third test after being diluted threefold with UTM™.

Finally, we compared the results of 814 nasal and 841 nasopharyngeal samples obtained in the Liat assay to those of the other molecular assays. The Liat assay was positive for SARS-CoV-2 in 113 (13.9%) of the nasal samples and 151 (18.0%) of the nasopharyngeal samples. The nasal sample was positive for influenza virus as detected by the Liat assay.

### 3.1 Comparison of the results of the Liat test with the results of other molecular assays using nasal samples

The results of the Liat assay are compared with the results of the cobas and NIID assays performed using nasal samples in Tables 1 and 2, respectively.

**Table 1.** Comparison of the results of the Liat and cobas SARS-CoV-2 assays performed using nasal samples.

|  |  | All cases |  | Symptomatic cases |  | Asymptomatic cases |  |
| --- | --- | --- | --- | --- | --- | --- | --- |
|  |  | Liat |  | Liat |  | Liat |  |
|  |  | Positive | Negative | Positive | Negative | Positive | Negative |
| cobas | Positive | 108 | 1 | 61 | 0 | 47 | 1 |
|  | Negative | 5 | 700 | 2 | 280 | 3 | 420 |
| Total concordance |  | 99.3 (98.4–99.7) |  | 99.4 (97.9–99.9) |  | 99.2 (97.8–99.8) |  |
| Positive concordance |  | 99.1 (94.5–100) |  | 100 (94.1–100) |  | 97.9 (88.9–99.9) |  |
| Negative concordance |  | 99.3 (98.4–99.8) |  | 99.3 (97.4–99.9) |  | 99.3 (97.9–99.9) |  |

cobas, cobas 6800 system and cobas SARS-CoV-2 & Influenza AB; Liat, cobas Liat system and cobas Liat SARS-CoV-2 & Influenza AB; SARS-CoV-2, severe acute coronavirus 2
The 95% confidence intervals are given in parentheses.

**Table 2.**
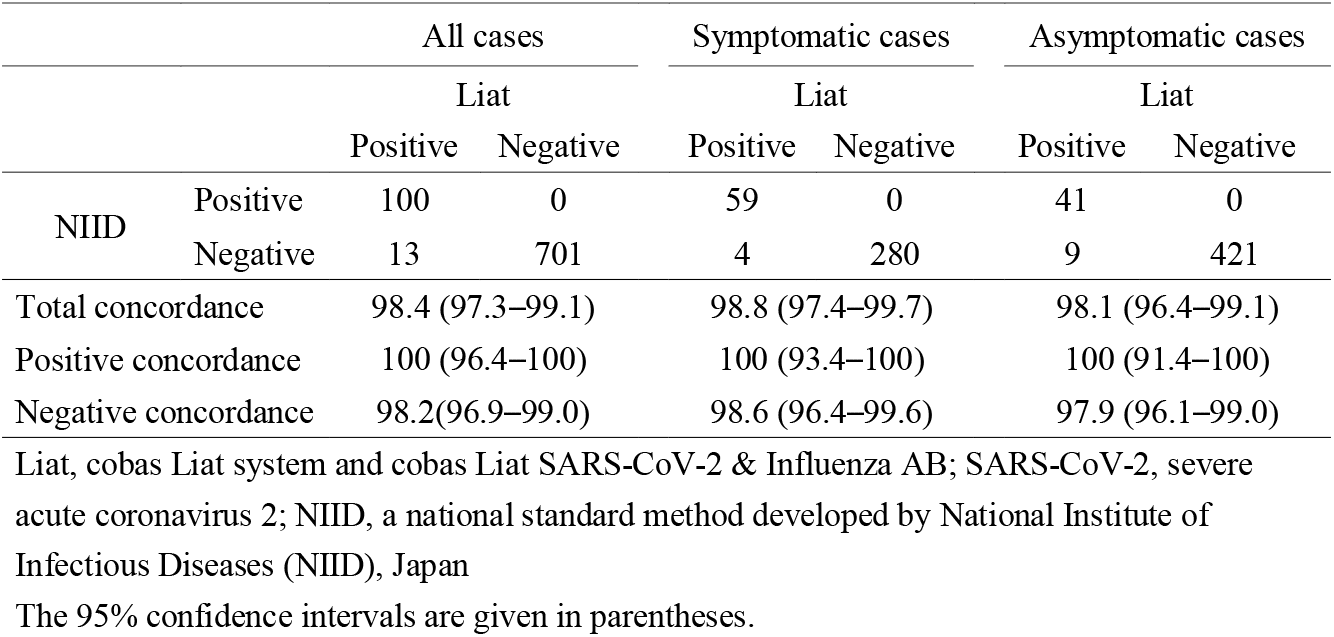
Comparison of the results of the Liat and NIID SARS-CoV-2 assays performed using nasal samples.

The total, positive, and negative concordance between the Liat and cobas assays were 99.3%, 99.1%, and 99.3%, respectively. The comparison was also performed stratified by the presence of relevant symptoms. In the symptomatic participants, the total, positive, and negative concordance was 99.4%, 100%, and 99.3%, respectively. In the asymptomatic participants, the total, positive, and negative concordance was 99.2%, 97.9%, and 99.3%, respectively.

Five nasal samples were positive only on the Liat assay, with Ct values ranging from 31.9 to 37.2. One of the five samples tested positive on additional analyses using Xpert Xpress with the Ct values of 39.6 for the E gene and 37.3 for the N2 gene.

When compared to the results of the NIID test, 13 sets of samples showed discordant results, all of which were Liat positive/NIID negative (Table 2). Eight of the 13 samples were positive on cobas assay. There were six sets of discordant results between the cobas and Liat assays, of which one sample was Liat-negative/cobas-positive and five samples were Liat positive/cobas negative.

### 3.2 Correlation of the cycle threshold values of the Liat assay with those of the other assays

The correlation of the Ct values of the Liat assay and the other assays is shown in Figure 1 (a, b, c). The median Ct values for each assay were as follows: Liat, 16.70; NIID, 23.7; cobas ORF1a/b gene, 24.4; cobas E gene, 23.6. The Ct values of the Liat assay were significantly lower (*p* < 0.001) but were correlated (*p* < 0.001) with those of the other assays.

**Fig. 1.**
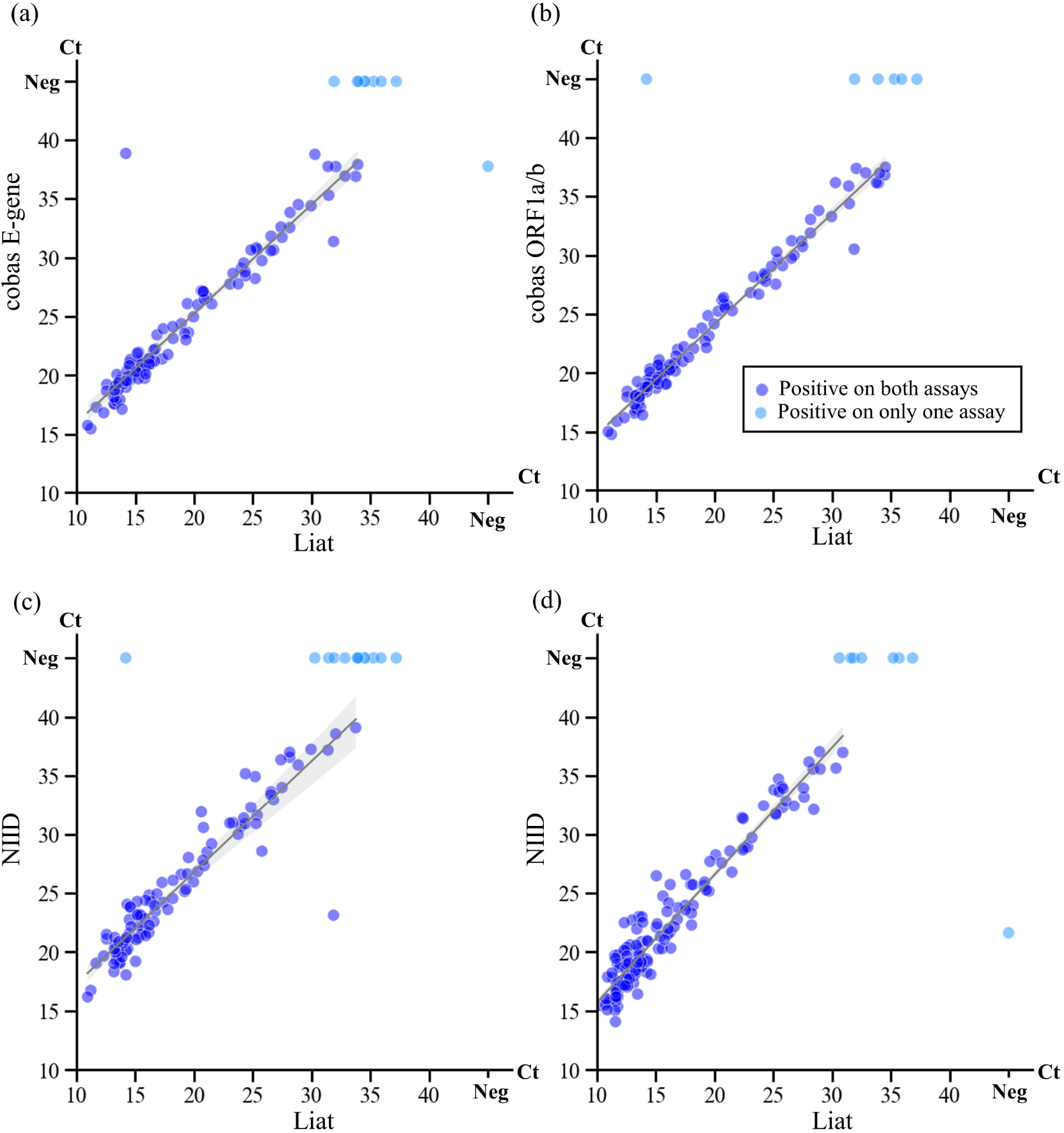
Comparison of the cycle threshold values of Liat with those of other molecular assays using the same samples. (a) Liat vs. cobas E-gene in nasal samples; (b) Liat vs. cobas ORF1a/b in nasal samples; (c) Liat vs. NIID in nasal samples; (d) Liat vs. NIID in nasopharygeal samples. The lines and surrounding gray areas indicate linear regression lines with 95% confidence intervals. The blue circles are samples for which both molecular assays tested positive. The light blue circles are samples for which one assay was positive and the other was negative. Abbreviations: cobas, cobas 6800 system and cobas SARS-CoV-2 & Influenza AB (cobas); Ct, cycle threshold; Liat, cobas Liat system and cobas Liat SARS-CoV-2 & Influenza AB; NIID, national standard method developed by the National Institute of Infectious Diseases of Japan

### 3.3 Analytical performance of the Liat assay using nasopharyngeal samples

Of the 841 nasopharyngeal samples included in the final analysis, the results of the Liat and NIID assays were positive for SARS-CoV-2 in 151 and 144 samples, respectively, with a concordance of 99.2%. There were seven discordant samples, all of which were Liat positive/NIID negative. The Ct values of the two assays were significantly correlated (*p* < 0.001, Fig. 1d).

Both nasopharyngeal and nasal samples were obtained from 813 participants. The test results of the Liat assay performed on the two sample types were compared. Both sample types tested positive in 108 samples, 14 pairs of samples tested positive on the nasopharyngeal sample and negative on the nasal sample, and four pairs of samples tested positive on the nasal sample and negative on the nasopharyngeal sample. The correlation of the Ct values between the two sample types is shown in Figure 2 (a, b).

**Fig. 2.**
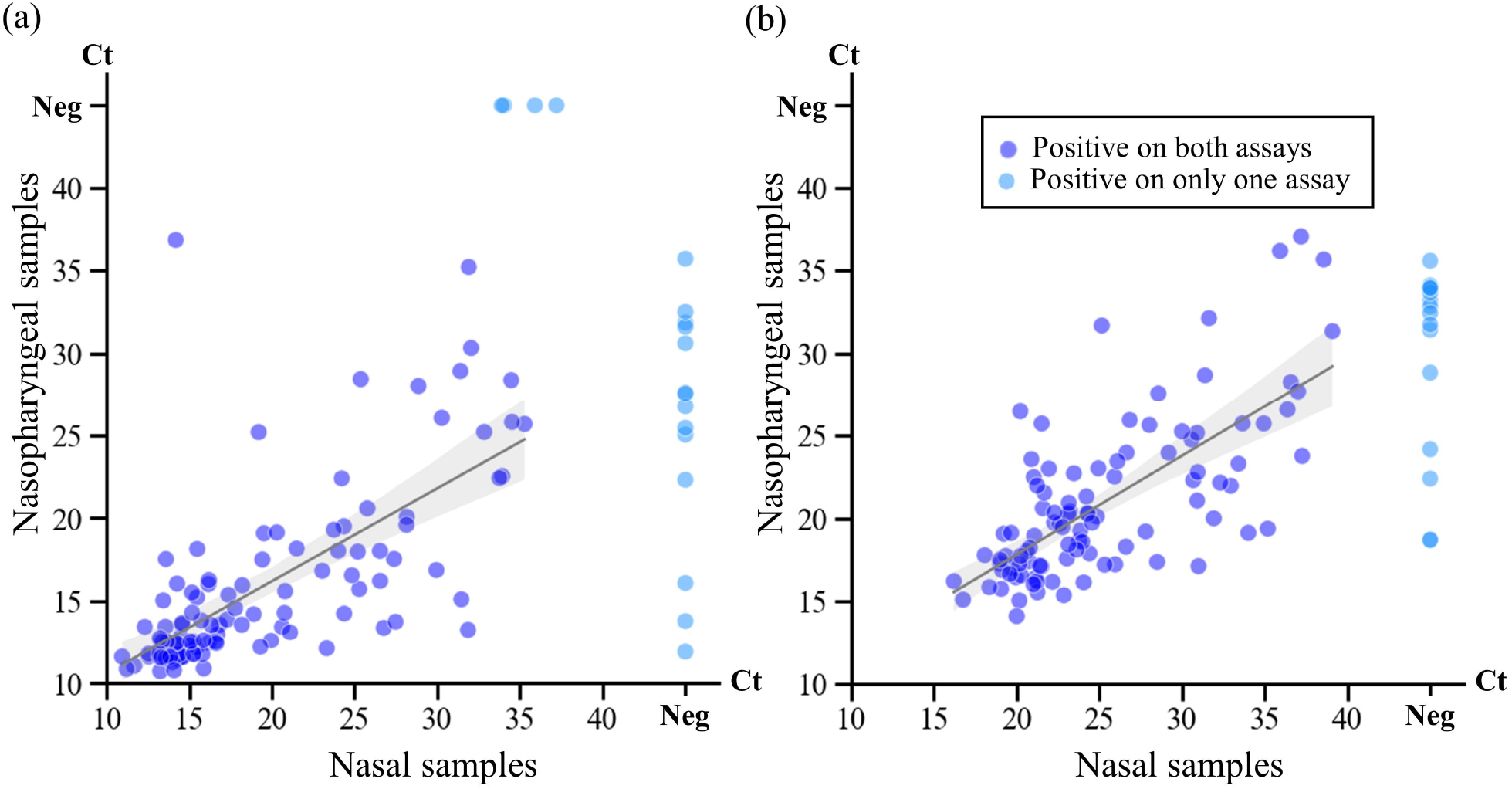
Comparison of cycle threshold values between the nasal and nasopharyngeal samples collected from same participants (a) Liat; (b) NIID. Abbreviations: cobas Liat system and cobas Liat SARS-CoV-2 & Influenza AB; Ct, cycle threshold; Liat, NIID, national standard method developed by the National Institute of Infectious Diseases of Japan

### 3.4 Testing of discordant samples using different molecular assays

Table 3 summarizes the results of the discordant cases: nasal samples with positive results on only one of the three molecular assays (left panel) and nasopharyngeal samples with discordant results of the Liat and NIID assay (right panel). Additional analyses using Xpert Xpress were performed for these cases, although one sample (case ID: T0198) could not be tested due to the lack of residual sample volume.

**Table 3.** Cases with discordant results between molecular assyas.

| Case Number | Symptoms | Nasal samples |  |  |  | Nasopharyngeal samples |  |  |
| --- | --- | --- | --- | --- | --- | --- | --- | --- |
|  |  | Liat | NIID | cobas | Xpert Xpress | Liat | NIID | Xpert Xpress |
| T0161 | – | Neg | Neg | Neg | NA | Pos (35.7) | Neg | Pos (E: Neg, N2:42.0) |
| T0198 | – | Neg | Neg | Neg | NA | Pos (31.9) | Neg | NA |
| T0267 | – | Neg | Neg | Neg | NA | Pos (30.6) | Neg | Pos (E:34.8, N2:38.1) |
| T0339 | – | Pos (14.2) | Neg | Pos (38.8) | NA | Pos (36.8) | Neg | Pos (E: Neg, N2:41.6) |
| T0401 | – | Neg | Neg | Neg | NA | Pos (31.6) | Neg | Pos (E: Neg, N2:39.6) |
| T0438 | – | Neg | Neg | Neg | NA | Pos (32.5) | Neg | Pos (E:40.0, N2:42.1) |
| T0556 | – | Pos (35.3) | Neg | Neg | Neg | Pos (25.7) | Pos (34.1) | NA |
| T0625 | + | Pos (31.9) | Neg | Neg | Pos (E:39.6, N2:37.3) | Pos (35.2) | Neg | Pos (E: Neg, N2:41.7) |
| T0690 | + | Pos (37.2) | Neg | Neg | Neg | Neg | Neg | NA |
| T0735 | – | Neg | Neg | Pos (T2:37.8) | Neg | Pos (13.8) | Pos (18.7) | NA |
| T0760 | – | Pos (35.9) | Neg | Neg | Neg | Neg | Neg | NA |
| T0778 | – | Pos (33.9) | Neg | Neg | Neg | Neg | Neg | Neg |

Discordant cases are defined as follows: for nasal samples, cases with positive results on only one of the three molecular assays (Liat, NIID, and cobas); for nasopharyngeal samples, cases with discordant results of the Liat and NIID assay.
Liat, cobas Liat system and cobas Liat SARS-CoV-2 & Influenza AB; NA, not available; Neg, negative; NIID, a national standard method developed by National Institute of Infectious Diseases (NIID), Japan; Pos, positive; SARS-CoV-2, severe acute coronavirus 2; Xpert Xpress, Gene Xpert system and the Xpert Xpress SARS-CoV-2

In nasal samples, five samples were positive only using the Liat assay (Ct value range: 31.9–37.2), and one sample was positive only using the cobas assay (Ct value: 37.8). Of the five Liat-positive discordant samples, one sample tested positive using the Xpert Xpress assay, and another was positive on the corresponding nasopharyngeal sample from the same participant on both the Liat and NIID assays.

In the nasopharyngeal samples, all seven discordant samples were Liat positive and NIID negative (Ct value range: 30.6–36.8). The Xpert Xpress assay results were positive in six of the seven discordant samples.

## 4. Discussion

This prospective evaluation showed the high concordance in the nasal samples of the Liat assay results with those of other molecular assays. The Ct values of the Liat assay were significantly lower than those of other molecular assays, but were significantly correlated, although some discordant results were observed. Most of the cases of discordance occurred in samples with Ct values >30 on the Liat assay.

In nasopharyngeal samples, the total, positive and negative agreement of the results of the Liat and the cobas 6800/8800 assays were 98.6%, 100%, and 97.4%, respectively [8]. Similarly, this study showed high concordance between the results of the Liat assay and other molecular assays in nasal samples. The Liat assay can provide accurate results in 20 minutes and the equipment is compact. This enable the Liat system to be applied in a variety of settings, such as for rapid diagnosis in primary care facilities, screening and infection control in admitted patients, and the contact tracing in individuals with a close contact history.

Some discordant results were observed both in nasal and nasopharyngeal samples, the majority of which were Liat positive and NIID/cobas negative. The discordant samples generally had Ct values >30 on the Liat assay, indicating low viral concentrations. The Liat, cobas, and NIID assays have all been shown to have high analytical performance in previous studies[8,17,20]. However, the analytical performance on clinical specimens may be different due to the quality of the RNA extraction, the presence of inhibitors, genomic mutations, and stochasticity observed in samples with very low viral concentrations [21,22]. A previous study comparing Liat and cobas reported that all discordant samples were Liat positive/cobas negative [8], which is consistent with the results of this study.

The Ct values of the Liat assay were strongly correlated with those of the cobas and NIID assays, but were significantly lower. Determining the Ct values is crucial to identify which patients with SARS-CoV-2 infection are most likely to transmit the virus, with higher Ct values indicating lower infectivity [23,24]. However, Ct values can vary depending on the reagents and equipment used, even if the same samples are tested [24]; thus, the Ct values provided by each set of equipment should be carefully interpreted.

The study also compared the results of the tests of the nasal samples with those of the nasopharyngeal samples collected from the same participants. Some discordance occurred between nasal and nasopharyngeal RT-PCR results, with the nasal samples being more likely to provide negative results. A meta-analysis found that the sensitivity of RT-PCR of nasal samples was 82% compared to RT-PCR of nasopharyngeal samples [25]. In this study, 88.2% of nasal samples tested positive using the Liat assay among participants whose nasopharyngeal samples tested positive using the Liat assay. The Ct values between both sample types were strongly correlated but varied. The viral load is generally lower in the nostrils than in the nasopharynx [25], although the procedures for sample collection and the conditions of samples may also have caused the fluctuation of their Ct values [24].

In the coming winter season, the influenza virus is likely to co-circulate with SARS-CoV-2 infection [26,27]. Many clinical manifestations of influenza virus infection are similar to those of SARS-CoV-2 infection [28], which makes the clinical diagnosis difficult. Nevertheless, the distinction of the two viruses may affect the initial choice of treatment and infection control measure. The Liat system is reported to accurately identify the influenza virus [9], although due to the limited scope of the present study we did not verify the results by using other molecular assays. We consider that the system can make significant contributions to daily clinical practices because of its feature of detecting both viruses simultaneously.

Our study has some limitations. First, we were unable to perform cobas testing on nasopharyngeal samples. The cobas assay has a high sensitivity and has been widely used worldwide [29,30]. The level of discordance may vary depending on the equipment used for comparison. Second, we did not evaluate performance of the assays in samples from individuals infected with SARS-CoV-2 variants with gene mutations. The emergence of new variants could affect the diagnostic performance of the test. Third, the nasal samples used for molecular examination were collected after acquiring those used for antigen testing. The viral load in the nasal samples may have reduced due to the order of the procedure, which may have led to the variations in results obtained in the molecular examination. Finally, we did not use fresh samples for the evaluation of molecular examinations. Although to a miniscule degree, the storage process involving freezing and thawing also reportedly affects the viral load in samples [31].

In conclusion, the results of the Liat assay showed a high concordance with those of the other molecular assays in both nasal and nasopharyngeal samples. The findings of our study suggests that the Liat assay is suitable for use in a variety of clinical situations, primarily where accurate detection of SARS-CoV-2 is necessary.

## Data Availability

All data produced in the present study are available upon reasonable request to the authors

## Acknowledgments

We thank Yoko Ueda, Mio Matsumoto, Masaomi Matsubayashi, Yumiko Tanaka, Mika Yaguchi, Naoko Minagawa, and the staff at the Tsukuba Medical Center Hospital for their intensive support in this study.

The author Yuki Adachi and the technical support staff at Roche Diagnostics laboratory performed the reference RT-PCR testing. The authenticity of the data obtained could be guaranteed by the fact that all equipment used for PCR examinations were fully automated and that authors Yusaku Akashi and H. Suzuki individually validated appropriate data handling under the supervision of the University of Tsukuba.

## Declarations

### Ethics Approval

The study was performed in accordance with the principles of the Declaration of Helsinki and with the STROBE guidelines and was approved by the ethics committee of the University of Tsukuba (approval number: R03-41).

### Consent to Participate

Verbal informed consent was obtained from all participants.

### Consent for Publication

Not applicable.

### Authors’ contributions

All authors meet the authorship criteria set by the International Committee of Medical Journal Editors. Yusaku Akashi was the principal investigator, wrote the first draft of the manuscript, and performed the statistical analyses. Michiko Horie, Kenichi Togashi, and Hiromichi Suzuki designed this study. Yuki Adachi and Junichi Kiyotaki performed the molecular testing. Shigeyuki Notake, Atsuo Ueda, and Koji Nakamura collected the samples and performed the diagnostic testing. Hiromichi Suzuki supervised the project. All authors contributed to writing the final draft of the manuscript.

